# Assessment of Clinical Competence Using AI-Standardized Clinical Examination

**DOI:** 10.64898/2026.10.01.26364500

**Authors:** Antonio Lopez, Ludovic Gabellier, Clément Peytavi, Zeno Loi, Emma Poupin-Lavigne, Catherine Plotton, Vincent Dupont, Léon Jahdauti, Philine de Vries, Philippe Thuillier, Maxence Compagnat, Pascale Vergne-Salle, Mathieu Kuchenbuch, Eva Feigerlova, Philippe Berthelot, Nathalie Bednarek, Béatrice Cochener-Lamard, Pierre-Yves Robert, Marc Braun, Denis Morin, Philippe Deruelle, Isabelle Laffont, Yves-Marie Pers, Kévin Yauy

**Author notes:** Corresponding authors: Antonio Lopez, MD,; Kévin Yauy, MD, PhD. These authors contributed equally (Antonio Lopez, Ludovic Gabellier).

## Abstract

**BACKGROUND:** As artificial intelligence (AI) enters clinical practice, concerns about deskilling and never-skilling make the independent assessment of clinical competence increasingly important. The AI-Standardized Clinical Examination (ASCE) uses AI to simulate and score standardized clinical encounters, potentially overcoming some of the logistical and standardization constraints of conventional objective structured clinical examinations (OSCEs). Whether it provides valid and reliable assessment at scale is unknown.

**METHODS:** In this prospective multicenter cohort study, final-year medical students from six French medical schools completed a 10-station ASCE six weeks before the national OSCE session. GPT-4o simulated standardized clinical partners; transcripts were subsequently scored by GPT-4.1 against faculty-defined rubrics. The primary outcome was the correlation between ASCE and national OSCE scores. Secondary outcomes were inter-rater reliability of expert transcript scoring, alignment of ASCE scores with expert ratings, and internal consistency.

**RESULTS:** Among 948 enrolled students, 588 had paired ASCE and national OSCE scores. ASCE scores were correlated with national OSCE performance (Spearman’s ρ, 0.57; 95% CI, 0.51 to 0.63), with less than 1% of residual variance attributable to center (intraclass correlation coefficient, 0.01). Eight experts scored 20 randomly selected ASCE transcripts; Krippendorff’s α was 0.77 overall, 0.80 for clinical skills, and 0.70 for communication and attitude skills. Adding GPT-4.1 as an evaluator reduced α by 0.02, and its scores did not differ significantly from those of individual experts. Internal consistency was 0.67 across 10 stations and was projected to approach 0.80 with 20 stations.

**CONCLUSIONS:** Scores from an examination delivered and scored by AI were associated with performance on a national high-stakes OSCE, with minimal center-level variation, tentative agreement among expert raters, and automated scores that aligned with expert ratings. These results support AI delivery and scoring of clinical examinations, with expert review of automated scores.

**Description:** In what is, to our knowledge, the first prospective multicenter study to evaluate a clinical examination delivered and scored by AI against an independent national OSCE, 588 final-year medical students across six French medical schools completed an AI-Standardized Clinical Examination before the independent national OSCE. Scores correlated with national OSCE performance (Spearman’s ρ, 0.57), with negligible variation between centers, supporting AI-standardized assessment at scale.

## INTRODUCTION

As artificial intelligence (AI) is integrated into clinical practice, concerns about deskilling or never-skilling and eroded clinical reasoning make the assessment of competence increasingly important [1,2]. Traditional time-based medical education produces variable outcomes and evidence suggests it does not consistently prepare learners for practice [3]. Competency-based medical education (CBME) addresses this by anchoring progression in the demonstrated mastery of explicitly defined competencies rather than in time served [3]. Objective structured clinical examinations (OSCEs) are a widely used method for assessing clinical skills. Introduced half a century ago to address the subjectivity of bedside examinations [4], OSCEs have since been adopted worldwide [5]. However, they remain resource-intensive, requiring recruited and trained standardized patients [6], dedicated facilities, and coordinated scheduling across multiple evaluators. Their reliability is also limited by the number of stations [7].

AI is increasingly used in clinical skills training and assessment [8]. AI-simulated patient systems have been evaluated for OSCE preparation, including nonrandomized and pilot randomized trials [9,10]. AI-based scoring of OSCE post-encounter notes and written clinical reports has also shown promising correlation with expert raters [11,12]. More recently, integrated systems have combined AI-simulated patients with automated assessment, validated against human ratings of the same transcripts [13,14]. Yet, whether scores from an AI-delivered and AI-scored clinical examination reflect performance on an independent high-stakes examination remains unknown [15]. We described an AI-standardized clinical examination (ASCE) that operationalizes core CBME principles while easing the logistic challenges of conventional OSCEs [16,17]. ASCE is a text-based examination in which large language models simulate standardized encounters and score the transcripts against faculty-written rubrics. In one randomized trial, ASCE-based preparation improved students’ subsequent OSCE performance [17]; in another, the ASCE served as the outcome measure for podcast-based preparation [18]. The French national OSCE offers an external criterion: a standardized, high-stakes examination that ranks students for specialty allocation rather than deciding pass or fail [19], independent of the ASCE and taken by the same students six weeks later. Our objective was to evaluate the validity and reliability of the ASCE as an assessment of clinical competence by determining whether ASCE scores correlate with national OSCE performance (criterion-related validity), whether ASCE transcripts could be scored reproducibly by expert physicians (inter-rater reliability), whether ASCE scores aligned with expert ratings, and how many stations are needed for reliable scores.

## METHODS

### STUDY DESIGN AND PARTICIPANTS

We conducted a multicenter prospective cohort study in six French medical schools: Montpellier-Nîmes (coordinating center), Brest, Limoges, Nancy, Reims, and Saint-Étienne (Figure 1).

**Figure 1:**
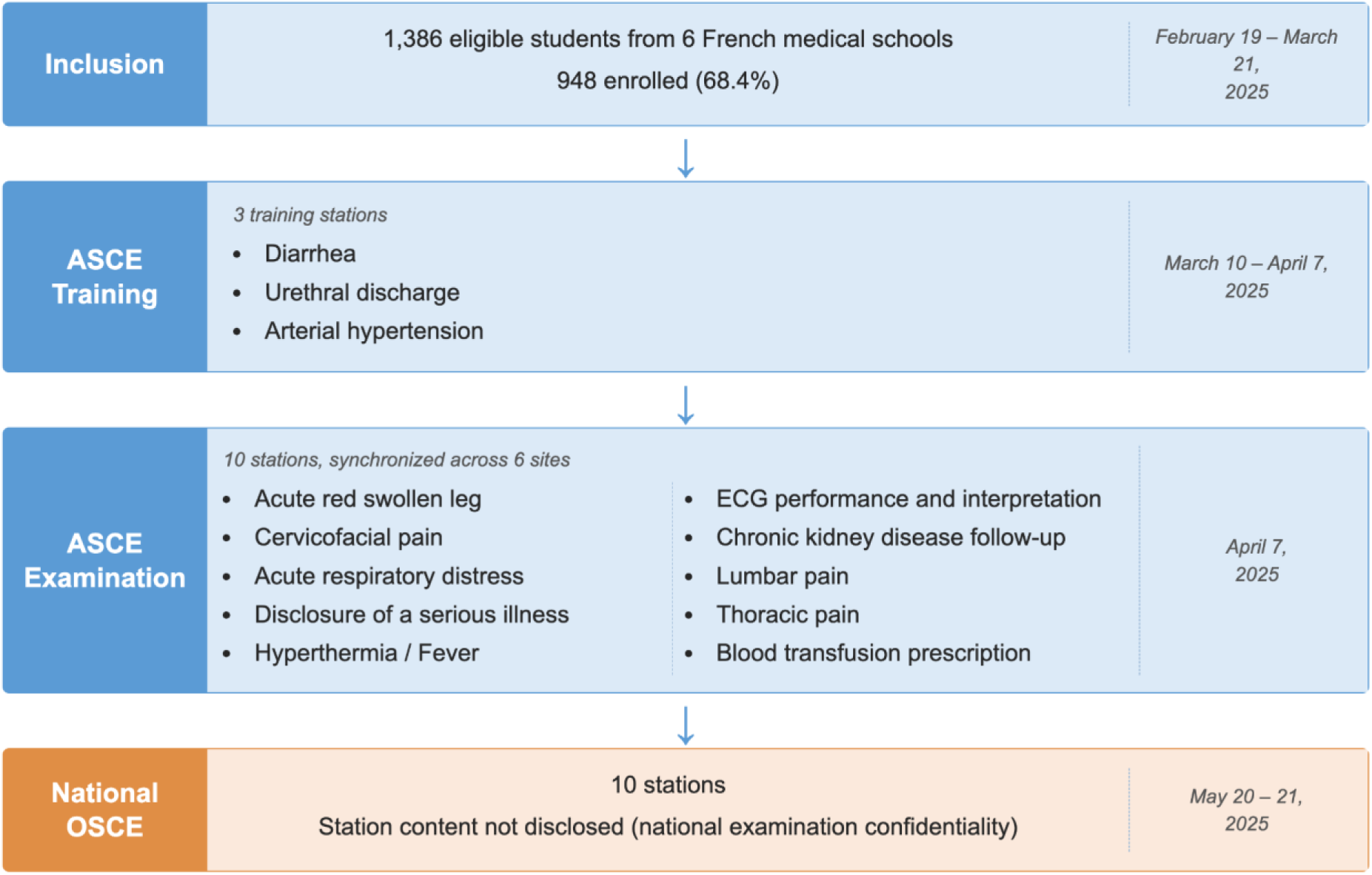
Study Protocol and Timeline. The study was conducted between February and May 2025 at six French medical schools. The inclusion period ran from February 19 to March 21, 2025. Three training stations were available from March 10 to April 7, 2025, before a 10-station ASCE session on April 7 (12 minutes per station, 2:00 to 6:00 p.m., synchronous across all sites). The national OSCE was administered on May 20 and 21, approximately 6 weeks after the ASCE session. ASCE denotes AI-standardized clinical examination, and OSCE objective structured clinical examination.

All sixth-year medical students enrolled in the national OSCE preparation program at participating sites were eligible. Students were invited through their faculty mailing lists to complete an online inclusion questionnaire between February 19 and March 21, 2025, with up to five reminders.

### ASCE PLATFORM AND SESSION DESIGN

The ASCE platform is a text-based clinical simulation system in which students interact in real time with AI-generated standardized partners. The platform and its educational framework have been described [17]. Standardized partner encounters were generated with GPT-4o; transcripts were subsequently scored with GPT-4.1. The simulation model was instructed to generate partner responses strictly within the scenario parameters and not to introduce clinical information not defined in the case, maintaining reproducibility across encounters. The AI scoring system applied the same rubric to the full conversation transcript, which was also the basis for subsequent human expert evaluation.

Prior to the formal assessment, students had access to 3 training stations, to familiarize themselves with the platform interface, the text-based interaction format and the scoring rubric. For both the training phase and the formal session, students connected remotely through a web browser, from a location of their choosing, and could initiate each successive station at their own pace within the designated timeframe.

The ASCE session consisted of 10 original stations of 12 minutes each within a 4-hour window, delivered synchronously across all six centers. The ASCE stations were designed by a multidisciplinary panel of clinical faculty from the participating centers to cover 9 of the 11 evaluation domains assessed in the national OSCE, excluding physical examination and procedural skills, which require in-person interaction (Figure S1). Each station was built around a clinical scenario that specified the clinical context (presenting complaint, relevant history, examination findings available on request), the partner role, a script of expected responses to student queries, and the scoring criteria. AI scoring used a standardized rubric of 15 items per station: 10 to 12 binary clinical items (scored 0 or 1) and 3 to 5 communication/attitude items on a 5-level ordinal scale (0, 0.25, 0.50, 0.75, or 1.0), with total scores converted to a 20-point scale (Table S1).

### EVALUATION OF ASCE TRANSCRIPTS BY EXPERTS AND AI

Two ASCE transcripts from each station, for a total of 20 transcripts, were randomly selected for review. Eight expert evaluators (H1 through H8), all co-authors and clinical teachers from participating centers with at least 5 years of experience in OSCE creation and evaluation, were asked to score each transcript independently, using the same rubric applied during automated scoring. Evaluators were blinded to one another’s ratings and to the automated scores.

To compare individual evaluator performance against a consensus reference, a leave-one-out procedure was used [20]: the reference was the median of the scores assigned by the remaining expert evaluators, computed both item by item and on the summed scores. For binary clinical items this median is equivalent to a majority vote; items on which the remaining raters were evenly split (median 0.5; 36 of 1650 comparisons, 2.2%) were excluded. Because each evaluator was removed from the consensus against which they were scored, this design avoided incorporation bias [21] and yielded a human benchmark for GPT-4.1 [22]. GPT-4.1 was not part of the expert pool and was therefore compared with the median of all experts who scored that transcript.

### OUTCOMES

The national OSCE, a high-stakes examination, occurred 6 weeks after the ASCE session, and comprised 10 clinical stations with live standardized patients. National OSCE scores for students included in this study were obtained from the examination authorities after official release in June 2025.

The primary outcome was the correlation between ASCE session scores and national OSCE scores (criterion-related validity).

Three secondary outcomes were defined a priori in the protocol approved on January 31, 2025: (i) whether ASCE transcripts could be scored reproducibly by expert physicians (inter-rater reliability), (ii) whether AI-generated ASCE scores aligned with expert ratings (AI scoring agreement), and (iii) the minimum number of stations required for reliable assessment (internal consistency reliability). Consistency of the primary association across centers was a prespecified robustness analysis.

### STATISTICAL ANALYSIS

Among students who completed all 10 stations, the correlation between ASCE and OSCE scores (primary outcome) was assessed using Spearman rank correlation coefficients (ρ), computed overall and per center, with 95% confidence intervals from 1000 bootstrap replicates of student pairs; cross-site consistency was evaluated with an ASCE-by-center interaction test in a linear model. A mixed linear model was fitted with OSCE score as the dependent variable, ASCE score as the fixed effect, and center as a random intercept, yielding the marginal and conditional coefficient of determination (R²) [23] and the intraclass correlation coefficient. Inter-rater reliability (secondary outcome 1) was assessed using Krippendorff’s α for interval data, calculated separately for total, clinical, and communication/attitude scores, with 95% confidence intervals from 1000 bootstrap iterations. Values of α ≥ 0.80 were considered reliable, and values between 0.667 and 0.80 sufficient only for tentative conclusions (hereafter, tentative agreement) [24]. The same framework was applied after including GPT-4.1 in the evaluator pool (secondary outcome 2). Differences in scoring among evaluators were assessed on each evaluator’s signed deviation from a leave-one-out consensus, separately for each summed score. Because the Friedman test required complete blocks, it was restricted to the seven transcripts scored by all nine evaluators (eight experts and GPT-4.1); Wilcoxon signed-rank tests with Holm correction were applied for all available post-hoc pairwise comparisons.

For session reliability (secondary outcome 3), Cronbach’s α was computed across the ASCE stations, and the Spearman-Brown prophecy formula was used to project reliability at higher station counts.

To evaluate selection bias, national OSCE score distributions were compared with Welch analysis of variance among students who completed all ASCE stations, those who connected but did not complete, and enrolled students who did not participate.

No formal sample size calculation was performed; all eligible students at the six participating centers were invited during the inclusion period. All statistical tests were two-sided with a type I error of 0.05. All analyses were performed in Python, and figures were produced with R, version 4.5.2 (R Foundation for Statistical Computing).

### ETHICAL CONSIDERATIONS

This study was approved by the Montpellier University Research Ethics Committee (No. UM 2025-011, January 31, 2025). All participants provided electronic informed consent. Student data were pseudonymized before analysis.

## RESULTS

### PARTICIPANTS AND ASCE SESSION

Of 1386 eligible sixth-year medical students, 948 (68.4%) were enrolled (inclusion cohort). On April 7, 2025, the formal ASCE session was administered synchronously across all six centers; 648 participated in the ASCE session (participant cohort), and 589 completed all 10 stations (completion cohort; 90.9% of participants and 62.1% of enrolled students) (Figure 2). Compared with the 300 enrolled students who did not participate, participants were more often women (71% vs. 57%) and more often reported digital-platform training (63% vs. 49%); other differences were smaller (Table 1). The national OSCE took place on May 20 and 21, 2025. Paired ASCE and national OSCE scores were available for 588 students (primary analysis cohort). Among the 937 enrolled students with an available national OSCE score, scores did not differ between those who completed all ASCE stations (n = 588), those who connected but did not complete (n = 56), and those who did not participate (n = 293) (Figure S2); the 11 remaining enrolled students had no national score.

**Figure 2:**
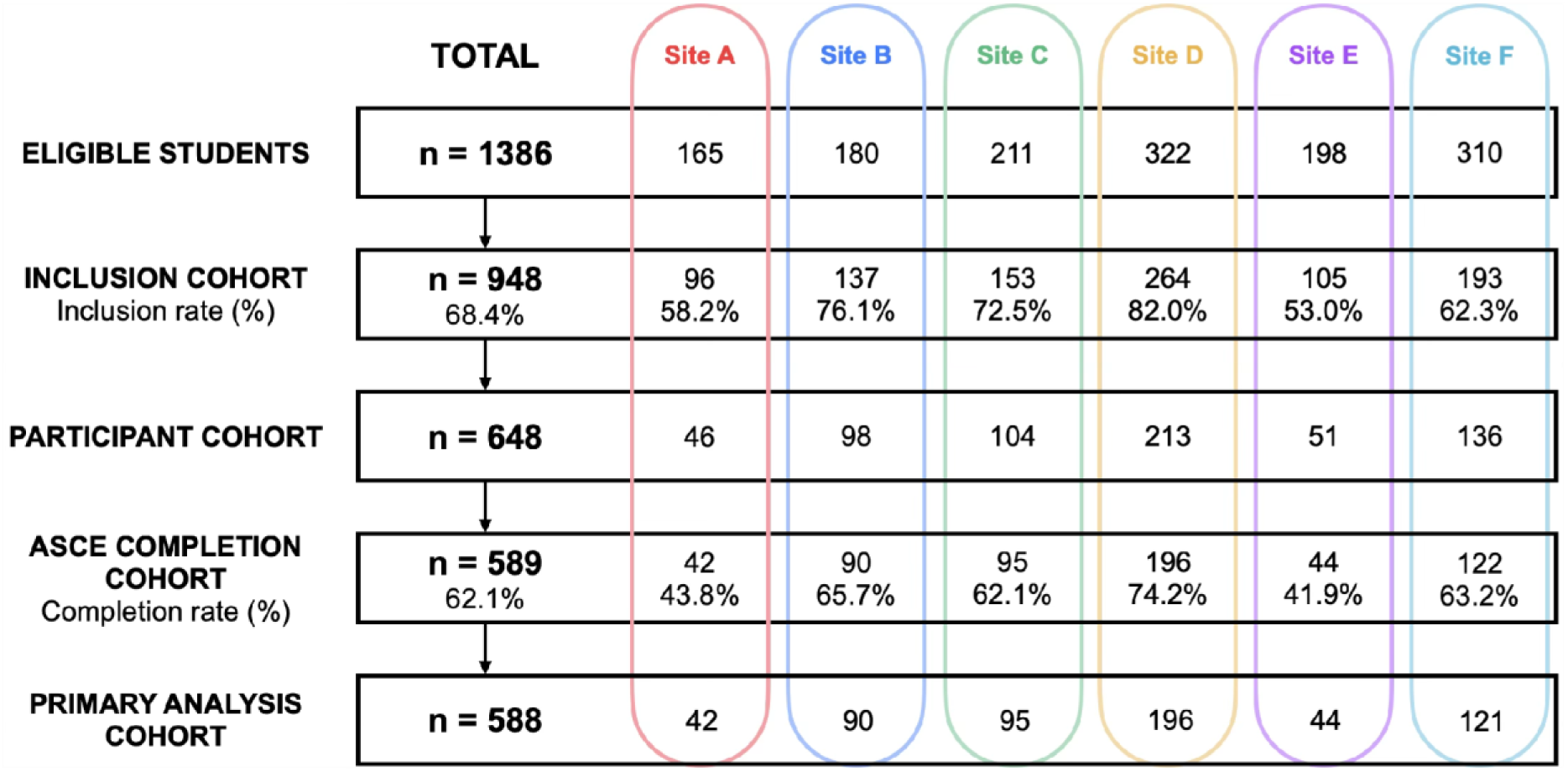
Participant Flowchart. Of 1386 eligible sixth-year medical students across six centers, 948 (68.4%) were enrolled, 648 participated in the ASCE session, 589 completed all 10 stations (90.9% of participants and 62.1% of enrolled students), and 588 had paired ASCE and national OSCE scores for the primary analysis. Center-level enrollment and completion numbers are shown. ASCE denotes AI-standardized clinical examination, and OSCE objective structured clinical examination.

**Table 1:** Baseline Characteristics of the Study Population. Values are reported as n (%) for categorical variables and mean ± SD for continuous variables. The inclusion cohort comprises all enrolled students, the participant cohort those who began the AI-standardized clinical examination session, and the completion cohort those who completed all 10 stations. Percentages are calculated on the number of respondents for each item, which may be lower than the cohort size shown in the column header. The three cohorts are nested: the participant cohort is a subset of the inclusion cohort, and the completion cohort a subset of the participant cohort. Counts for enrolled students who did not participate are obtained by subtracting the participant column from the inclusion column; the corresponding proportions are computed on the difference of the item-level denominators.

| Characteristic | Inclusion Cohort (n=948) | Participant Cohort (n=648) | Completion Cohort (n=589) |
| --- | --- | --- | --- |
| **Female sex — n (%)** | 625 (66.7) | 457 (71.2) | 415 (71.1) |
| **Age — mean (yr) ± SD** | 24.3 ± 2.3 | 24.4 ± 2.4 | 24.4 ± 2.4 |
| **Honors on the baccalauréat\* — n (%)** |  |  |  |
| With honors | 482 (50.9) | 344 (53.1) | 308 (52.3) |
| Good | 294 (31.1) | 202 (31.2) | 186 (31.6) |
| Fairly good | 118 (12.5) | 70 (10.8) | 63 (10.7) |
| No honors | 27 (2.9) | 18 (2.8) | 18 (3.1) |
| Prefer not to say | 25 (2.6) | 14 (2.2) | 14 (2.4) |
| **Admission to medical studies via a bridging program — n (%)** | 70 (7.4) | 53 (8.2) | 47 (8.0) |
| **Total clinical rotations — mean ± SD** |  |  |  |
| In medical (non-surgical) services | 9.0 ± 3.2 | 9.1 ± 3.4 | 9.1 ± 3.4 |
| In surgical services | 2.2 ± 1.5 | 2.1 ± 1.4 | 2.1 ± 1.4 |
| **Repeated ≥ 1 year of medical school — n (%)** | 474 (50.1) | 326 (50.3) | 295 (50.1) |
| **Personal issues during studies — n (%)** |  |  |  |
| Yes | 356 (37.6) | 235 (36.3) | 212 (36.0) |
| No | 499 (52.7) | 354 (54.6) | 327 (55.5) |
| Prefer not to say | 91 (9.6) | 59 (9.1) | 50 (8.5) |
| **Concurrent employment — n (%)** |  |  |  |
| Yes | 312 (33.0) | 202 (31.2) | 181 (30.7) |
| No | 595 (62.9) | 422 (65.1) | 385 (65.4) |
| Prefer not to say | 39 (4.1) | 24 (3.7) | 23 (3.9) |
| **Faculty-led OSCE training — n (%)** | 561 (59.3) | 393 (60.6) | 359 (61.0) |
| **Modality of OSCE training — n (%)** |  |  |  |
| During clinical rotations | 841 (88.9) | 578 (89.2) | 525 (89.1) |
| Faculty-led peer-tutoring | 639 (67.5) | 449 (69.3) | 411 (69.8) |
| Private organizations | 283 (29.9) | 202 (31.2) | 180 (30.6) |
| Self-training (alone or with peers) | 817 (86.3) | 572 (88.3) | 523 (88.8) |
| Digital platform | 553 (58.5) | 408 (63.0) | 368 (62.5) |

\* Baccalauréat is equivalent to high school diploma.
SD denotes standard deviation, and OSCE objective structured clinical examination.

### ASCE SCORE CORRELATION WITH NATIONAL OSCE PERFORMANCE

Among the 588 students with paired scores, median scores were 10.97 (interquartile range [IQR], 9.93 to 11.83) for the ASCE and 14.36 (IQR, 13.41 to 15.31) for the national OSCE on a 20-point scale (Figure S3). ASCE scores correlated with national OSCE scores (Spearman’s ρ, 0.57; 95% confidence interval [CI], 0.51 to 0.63; Figure 3). In the mixed linear model, ASCE scores explained 34% of the variance in national OSCE scores (marginal R², 0.34; conditional R², 0.35). This correlation was consistent across the six centers (ρ range, 0.46 to 0.64), and between-center differences accounted for less than 1% of the residual variance in national OSCE scores after adjustment for ASCE performance (intraclass correlation coefficient, 0.01). Station-level ASCE mean scores are reported in Table S2.

**Figure 3:**
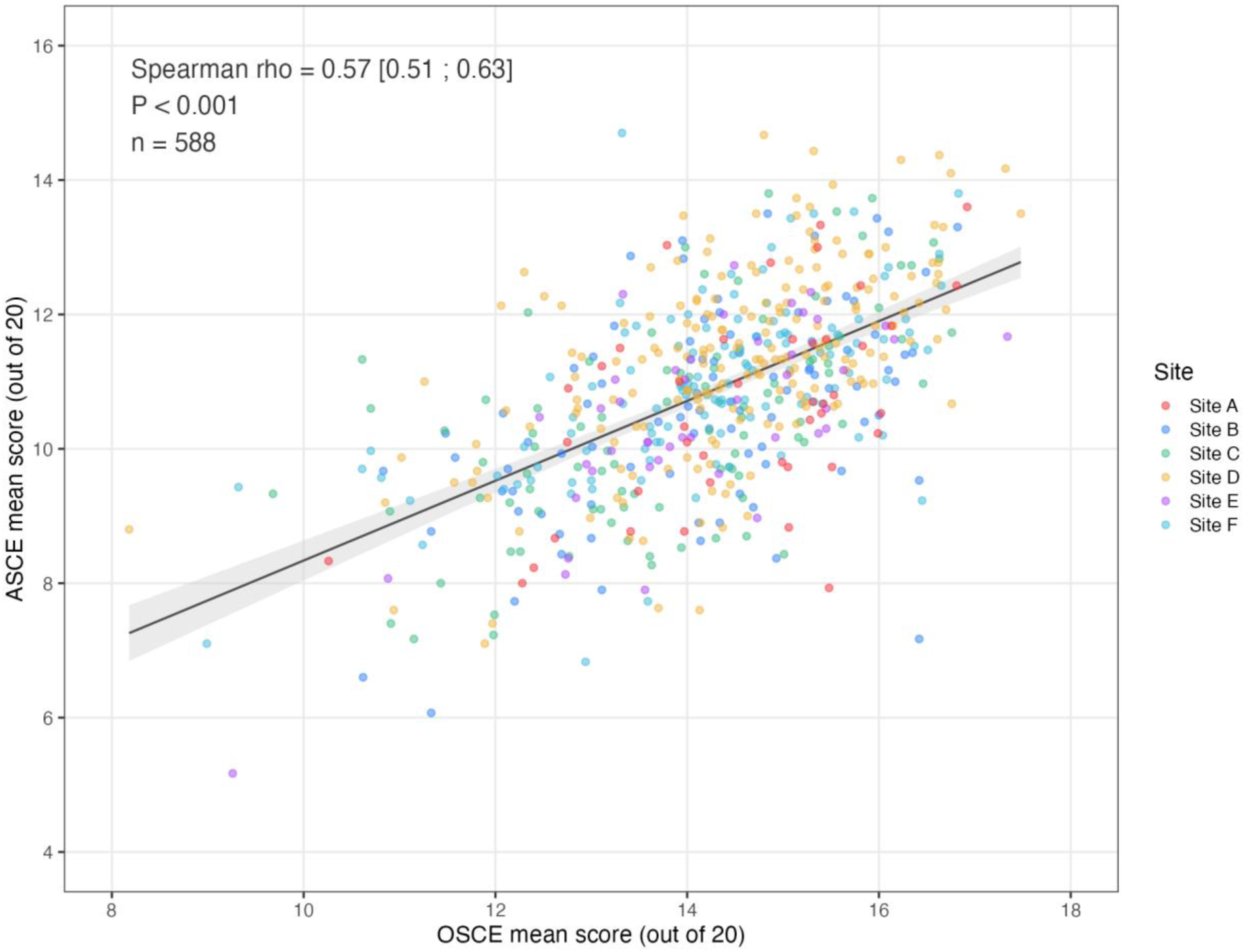
Correlation Between ASCE and OSCE Scores. Data are mean scores over 10 stations in ASCE and OSCE for 588 medical students. The solid line indicates the linear trend, and the shaded band its confidence interval; points are colored by site (A to F). ASCE and national OSCE rankings were positively associated (Spearman’s ρ, 0.57; 95% CI, 0.51 to 0.63). ASCE denotes AI-standardized clinical examination, and OSCE objective structured clinical examination.

### ASCE TRANSCRIPT SCORING REPRODUCIBILITY ACROSS EXPERT PHYSICIANS

Eight expert evaluators independently scored 20 ASCE transcripts, yielding 143 of 160 possible evaluations (89.4%); 17 were missing because some evaluators did not score all 20 transcripts. Krippendorff’s α was 0.77 (95% CI, 0.59 to 0.86) for total scores, 0.80 (95% CI, 0.61 to 0.89) for clinical skills, and 0.70 (95% CI, 0.22 to 0.85) for communication and attitude skills. Subdomain scoring patterns are shown in Figures S4 and S5.

### ASCE AUTOMATED SCORING AGREEMENT WITH EXPERT RATINGS

With GPT-4.1’s 20 scores added to the 143 expert evaluations, 163 scores on 20 transcripts were analyzed. Krippendorff’s α for total scores was 0.75 (95% CI, 0.56 to 0.86). For clinical skills, α was 0.78 (95% CI, 0.60 to 0.87); for communication and attitude, 0.68 (95% CI, 0.21 to 0.83). Adding GPT-4.1 reduced α by 0.02 across all three domains; the AI-inclusive α fell within the 95% confidence interval of the experts-only estimate.

Among the seven transcripts scored by all nine evaluators, the Friedman test detected evaluator differences for total scores (P = 0.01) and for communication and attitude (P = 0.02). In post-hoc comparisons using all transcripts available for each pair (36 evaluator pairs, Holm-corrected), GPT-4.1 did not differ significantly from any expert (all adjusted P > 0.07); the only significant difference was between two experts (adjusted P = 0.009). GPT-4.1’s deviations from the consensus were centered near zero but more dispersed than those of most experts (Figure 4).

**Figure 4:**
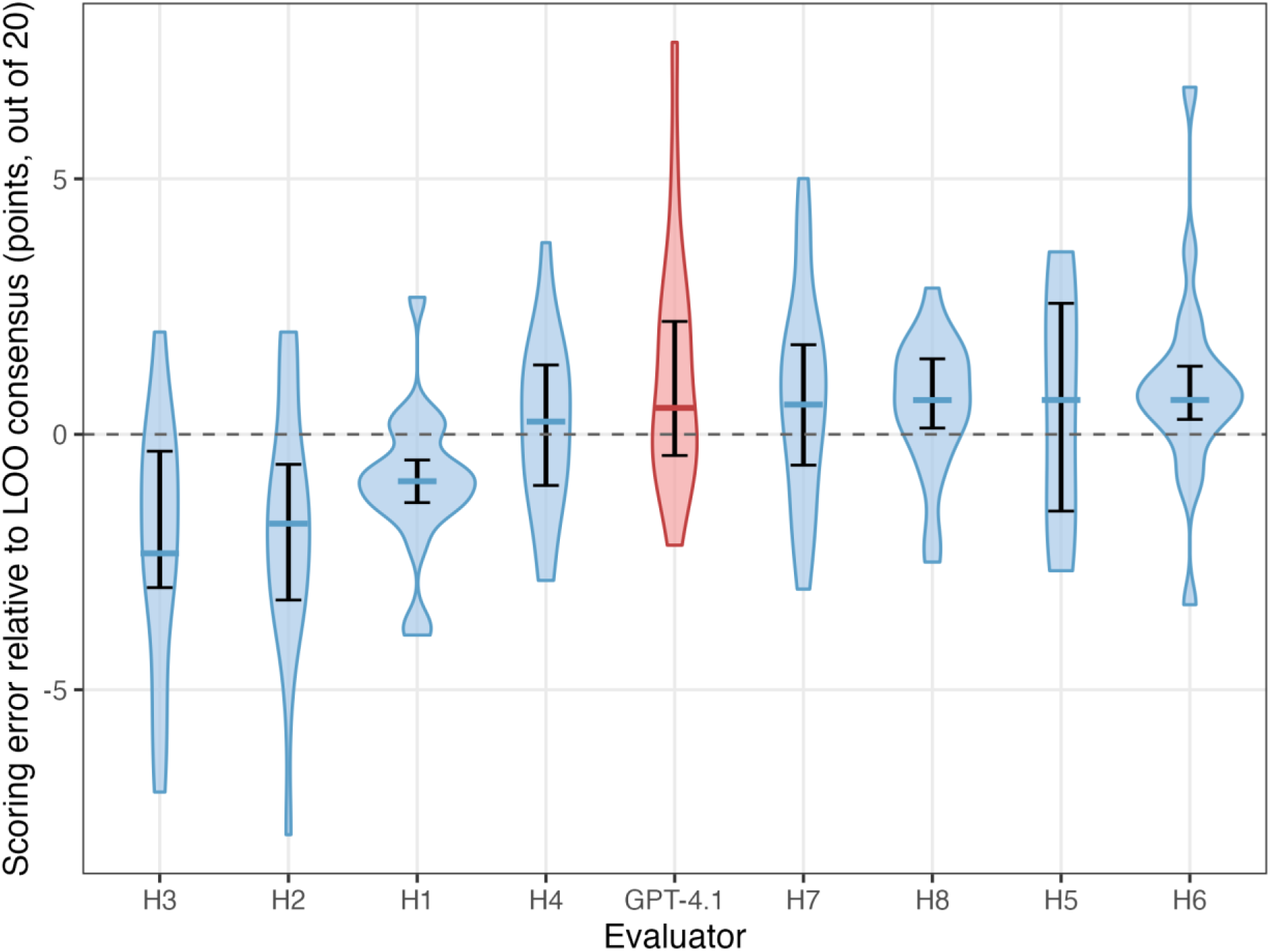
Comparative Scoring Error of Human Experts Versus AI. Violin plots displaying the distribution of scoring errors for eight human evaluators (H1 through H8) and the GPT-4.1 model. Error is the difference between an evaluator’s total score for a transcript and a leave-one-out expert consensus, defined as the median of the total scores assigned by the remaining expert evaluators; GPT-4.1 was compared with the median of all experts who scored that transcript. The analysis comprises 163 evaluations of 20 transcripts drawn from the 10 ASCE stations (2 per station). The width of each violin corresponds to the frequency of data points at that error magnitude. Evaluators are ordered by increasing median error. ASCE denotes AI-standardized clinical examination.

### ASCE INTERNAL CONSISTENCY

Across the 10 ASCE stations, Cronbach’s α was 0.67. Spearman-Brown projections indicated that approximately 20 stations would be required to reach α ≈ 0.80, and 45 for α ≈ 0.90 (Figure S6).

## DISCUSSION

In this multicenter prospective cohort of 588 final-year medical students, scores on a text-based examination in which one language model simulated standardized encounters and a second scored the transcripts were correlated with national OSCE performance (Spearman’s ρ, 0.57), with negligible residual between-center variance in OSCE scores after adjustment for ASCE performance (intraclass correlation coefficient, 0.01). Transcript scoring showed tentative agreement across eight independent expert physicians (Krippendorff’s α, 0.77) and was essentially unchanged when GPT-4.1 was added as a rater (α, 0.75); internal consistency was moderate at 10 stations (Cronbach’s α, 0.67).

To our knowledge, this is the first multicenter study to provide validity evidence based on relations to other variables for an examination both delivered and scored by AI against the national OSCE. Within the five sources of validity evidence [25], these data bear on content, internal structure, relations to other variables, and — through the accuracy of the automated scoring — response process; consequences were not evaluated. The moderate correlation is best read against its ceiling. Being text-based, the ASCE covers 9 of the 11 national OSCE domains but neither physical examination nor procedural skills, so perfect concordance would not be expected. In addition, OSCE scores are themselves imperfectly reliable, with a pooled across-station reliability of approximately 0.66 reported in a systematic review [7]. Measurement error in both examinations would therefore cap the observable correlation near 0.66 (√[0.67 × 0.66]) rather than 1 [26]. ASCE performance still explained 34% of the variance in national OSCE scores.

Residual between-center variance in OSCE scores was negligible. In conventional OSCEs, site, examiner, and standardized-patient differences add measurement error [27,28], and allocation to different examiner cohorts introduces systematic variance that conventional reliability indices do not detect [29]. Delivering and scoring every encounter with the same models and rubric is designed to remove examiner-related variance, although our data cannot separate this from a possible harmonization of national scores across centers.

Automated scoring is what makes cohort-scale standardization feasible. Adding GPT-4.1 to the rater pool lowered Krippendorff’s α by only 0.02 and produced no significant difference from any individual expert, consistent with prior reports of high model-human agreement on content items [11,30], significantly lower for communication items [13]. This is not equivalence. Automated evaluation of learning outcomes is high-risk under the European Union’s Artificial Intelligence Act (Annex III, point 3(b)) [31], which mandates effective human oversight from December 2027 (Article 14). Rather than reinstating exhaustive review, we propose a risk-targeted human-in-the-loop: expert rescoring of borderline or contested performances and of transcripts flagged as internally inconsistent or deviating markedly from the station distribution.

This study has several limitations. First, we obtained only the final national scores, without item-level data, and could therefore neither estimate the reliability of the national examination nor determine whether the scores had been harmonized across centers; the higher median of national OSCE scores (14.36 vs. 10.97) is consistent with such calibration. Second, the evidence that automated scoring can stand in for expert review remains preliminary: it rested on only 20 transcripts, with 17 of 160 expert ratings missing and wide confidence intervals for inter-rater reliability, particularly for communication/attitude scores; pooling transcripts across stations of unequal difficulty (Table S2) may also have inflated α. No equivalence margin was prespecified, the model’s errors, although centered near zero, were more dispersed than those of most experts, and the expert consensus criterion was itself only moderately reliable. Moreover, the encounters that experts rated were themselves generated by the simulation model, so that expert–model agreement bears on the scoring of a transcript, not on the fidelity of the simulated encounter. Concordance with this consensus therefore cannot demonstrate equivalence; an agreement coefficient modeling the reference group’s heterogeneity [32] would provide a convergent check. A prospective noninferiority design with a predefined margin, a larger transcript set, and an independently generated criterion would provide stronger evidence. Third, because the ASCE preceded the national examination by six weeks and ASCE preparation improves subsequent OSCE performance [17], a training effect could inflate the association; two features argue against it. Every included student had access to the same three training stations, an exposure too narrow to reproduce the previously reported usage gradient, so it would mostly shift the group mean. The coordinating center, whose students alone had used the platform for three months, showed no divergence from the other centers. Fourth, the ASCE is not yet suitable for high-stakes assessment: its ten-station internal consistency remains below the conventional threshold, and its text-based format cannot assess physical examination or procedural skills. A hybrid format combining an expanded ASCE with a few in-person stations could address both limitations. The ASCE was also delivered without proctoring, and unsupervised online assessment is open to misuse when participation is incentivized [33]. Fifth, communication and attitude items were scored less reliably than clinical items by both human and automated raters, consistent with across-station OSCE reliability [7] and with AI-based OSCE assessment [13]; a multimodal model scoring nonverbal behavior from video disagreed systematically with human raters (Krippendorff’s α, −0.47 [34]), so the nearer-term gain lies in redesigning communication rubrics for automated scoring rather than adding video. Sixth, all AI functions relied on proprietary, version-dependent models prompted zero-shot in one language, one country, and a single examination cycle. Each transcript was scored once, so run-to-run variability was not assessed, and silent model updates or deprecation threaten long-term reproducibility. Finally, only 42% of eligible students contributed to paired scores. Although national OSCE performance did not differ between participants and enrolled non-participants, non-enrolled students could not be compared, and voluntary participation favored women and students already training on digital platforms (Table 1). The absence of a formal sample size calculation and the proprietary rubrics and software also limit external replication.

As AI absorbs more clinical work, verifying that clinicians can reason and communicate independently becomes more important, not less. The decline in adenoma detection among endoscopists after routine AI exposure suggests that deskilling may already be occurring [35]. Addressing it requires assessing competence at scale.

These findings support AI delivery and scoring of standardized assessment, with expert judgment kept for review. The ASCE was developed and evaluated within DocSimulator and has since been integrated into Aptitudes, its university-operated successor (https://aptitudes.eu/), which uses open-weight models. With open-weight models, a given model version can be archived and the scoring rerun by independent groups, and successive model generations can be compared on the same archived transcripts. Text-based stations could cover the domains they assess, leaving in-person stations for physical examination, procedural skills, and interpersonal communication. The same technology that may contribute to deskilling can also help verify competence, if assessment stays transparent and experts review flagged transcripts.

## Data Availability

Deidentified participant data and analysis code may be made available upon reasonable request to the corresponding authors, subject to approval by the Montpellier University Research Ethics Committee and compliance with French data protection regulations.

## ACKNOWLEDGMENT

This work was funded by Université de Montpellier Programme d’Excellence I-SITE (2023, 2024, and 2026), Pôle Universitaire d’Innovation: Booster Innovation Montpellier (2024), and Région Occitanie. Emmanuel Guenou (compuute.io) contributed to the platform’s software development.

We thank Hervé Maisonneuve, MD, for critical review of the manuscript.

We thank the administrative and educational staff of the participating medical schools, Charline Duarte, Johanna Pletty, Laurent Truillet, Mathilde Raguin, Nolwenn Pommelet, Alexandra Lasnier, Magali Barry, Cyril Kahfujian, Charly Robert, Marion Berguer, Hind Hani, Thomas Thieblemont, Sébastien Cousin, and Maha Badreddine, for their help with student recruitment, examination logistics, and the collection of national OSCE scores.

## AI-Assisted Technology Disclosure

The DocSimulator platform (compuute.io) uses OpenAI language models to simulate standardized patient encounters and to score student transcripts. GPT-4o generated all simulated partner responses during the ASCE session of April 7, 2025. Transcripts were archived and subsequently scored against predefined rubrics by GPT-4.1, released on April 14, 2025. Both models were accessed through the OpenAI API during the study period (February to May 2025), and each transcript was scored in a single run. Clinical faculty designed the stations and the scoring rubrics, and a subset of 20 transcripts was independently rescored by eight expert physicians who were blinded to the automated scores. AI-assisted writing tools were used during manuscript preparation; all content was reviewed and edited by the authors, who take full responsibility for the accuracy and integrity of the work.

