## Supplementary material for "Assessment of Clinical Competence Using AI-Standardized Clinical Examination": Supp files

**Table of Contents**

Table S1. Description of ASCE Stations

Table S2. Station-Level Mean Scores

Figure S1. OSCE Assessment Domains Covered by ASCE

Figure S2. Selection Bias Analysis

Figure S3. Distribution of ASCE and OSCE Scores

Figure S4. Clinical Skills Scoring by Evaluator

Figure S5. Communication and Attitude Scoring by Evaluator

Figure S6. Predicted Reliability of ASCE

**Table S1. Description of ASCE Stations.**

The AI-standardized clinical examination (ASCE) comprised 10 examination stations (E1–E10), preceded by 3 training stations (T1–T3), all designed by a multidisciplinary panel of clinical faculty from the six participating centers. Training stations served to familiarize students with the platform; training performance was not included in the analysis. The 10 examination stations covered 9 of the 11 domains assessed in the national objective structured clinical examination (OSCE), excluding physical examination and procedural skills. Each station lasted 12 minutes and was scored on a 15-point scale (10 to 12 binary clinical items and 3 to 5 communication and attitude items), with total scores converted to a 20-point scale. The 10 examination encounters comprised 4 simulated patient consultations, 4 simulated nurse interactions, and 2 simulated parent consultations across medical and surgical specialties.

| Station | Encounter Type | OSCE Assessment Domain(s) | Initial Scenario | Main Item |
|---|---|---|---|---|
| **Training** | | | | |
| T1 | Simulated patient | Interview; Summary of paraclinical examination results | Diarrhea | Chronic diarrhea in adults and children |
| T2 | Simulated patient | Relevant management strategy; Education/Prevention | Urethral discharge | Sexually transmitted infections |
| T3 | Simulated patient | Interview; Relevant management strategy | Arterial hypertension | Arterial hypertension in adults and children |
| **Examination** | | | | |
| E1 | Simulated nurse | Interprofessional communication; Relevant management strategy | Acute red swollen leg | Deep venous thrombosis and pulmonary embolism |
| E2 | Simulated parent | Relevant management strategy | Cervicofacial pain | Tonsillitis and rhinopharyngitis in children |
| E3 | Simulated nurse | Iconography; Diagnosis strategy | Acute respiratory distress | Pleural effusion |
| E4 | Simulated patient | Diagnosis disclosure | Disclosure of a serious illness | Tumors of the colon and rectum |
| E5 | Simulated parent | Diagnosis strategy | Hyperthermia/Fever | Prolonged fever |
| E6 | Simulated nurse | Summary of paraclinical examination results; Emergency | ECG performance and interpretation | Electrocardiogram indications and interpretation |
| E7 | Simulated patient | Education/Prevention | Chronic kidney disease follow-up | Chronic kidney disease in adults and children |
| E8 | Simulated patient | Interview | Lumbar pain | Rachialgia (spinal pain) |
| E9 | Simulated patient | Interview | Thoracic pain | Acute chest pain |
| E10 | Simulated nurse | Interprofessional communication; Emergency | Blood transfusion prescription | Labile blood components and transfusion safety |

**Table S2. Station-Level Mean Scores.**

Mean ASCE scores for each station among the 589 students who completed all 10 stations (completion cohort) and the 648 students who participated (participant cohort). Each station was scored out of 15 points (10 to 12 binary clinical items and 3 to 5 communication and attitude items); station and overall scores are shown converted to a 20-point scale, as in the main manuscript.

| Station | Mean Score ± SD (Completion Cohort, n = 589) | Mean Score ± SD (Participant Cohort) |
|---|---|---|
| E1 | 6.4 ± 2.2 | 6.4 ± 2.2 (n = 610) |
| E2 | 10.1 ± 2.7 | 10.1 ± 2.8 (n = 611) |
| E3 | 7.9 ± 3.4 | 7.9 ± 3.4 (n = 597) |
| E4 | 15.8 ± 3.8 | 15.7 ± 3.8 (n = 603) |
| E5 | 10.9 ± 3.0 | 10.9 ± 3.1 (n = 596) |
| E6 | 10.4 ± 2.7 | 10.3 ± 2.8 (n = 631) |
| E7 | 7.0 ± 2.6 | 7.0 ± 2.6 (n = 622) |
| E8 | 16.1 ± 2.6 | 16.1 ± 2.6 (n = 648) |
| E9 | 13.3 ± 2.6 | 13.2 ± 2.6 (n = 636) |
| E10 | 10.7 ± 3.2 | 10.6 ± 3.2 (n = 640) |
| **Overall** | **10.87** ± 1.47 | — |

Scores are shown on a 20-point scale, per station and overall. For the participant cohort, the number of students evaluated at each station is provided; not all participants completed every station. ASCE denotes AI-standardized clinical examination, and SD standard deviation.

**Figure S1. OSCE Assessment Domains Covered by ASCE.**

Diagram showing the 11 assessment domains of the national objective structured clinical examination (OSCE). Nine domains (dark segments, green ring) were assessed by the AI-standardized clinical examination (ASCE): interview, diagnosis disclosure, relevant management strategy, diagnosis strategy, iconography, summary of paraclinical examination results, emergency, education/prevention, and interprofessional communication. Two domains (light segments, pink ring) were not assessed: physical examination and procedural skills, which require in-person interaction not supported by the text-based platform. ASCE denotes AI-standardized clinical examination, and OSCE objective structured clinical examination.

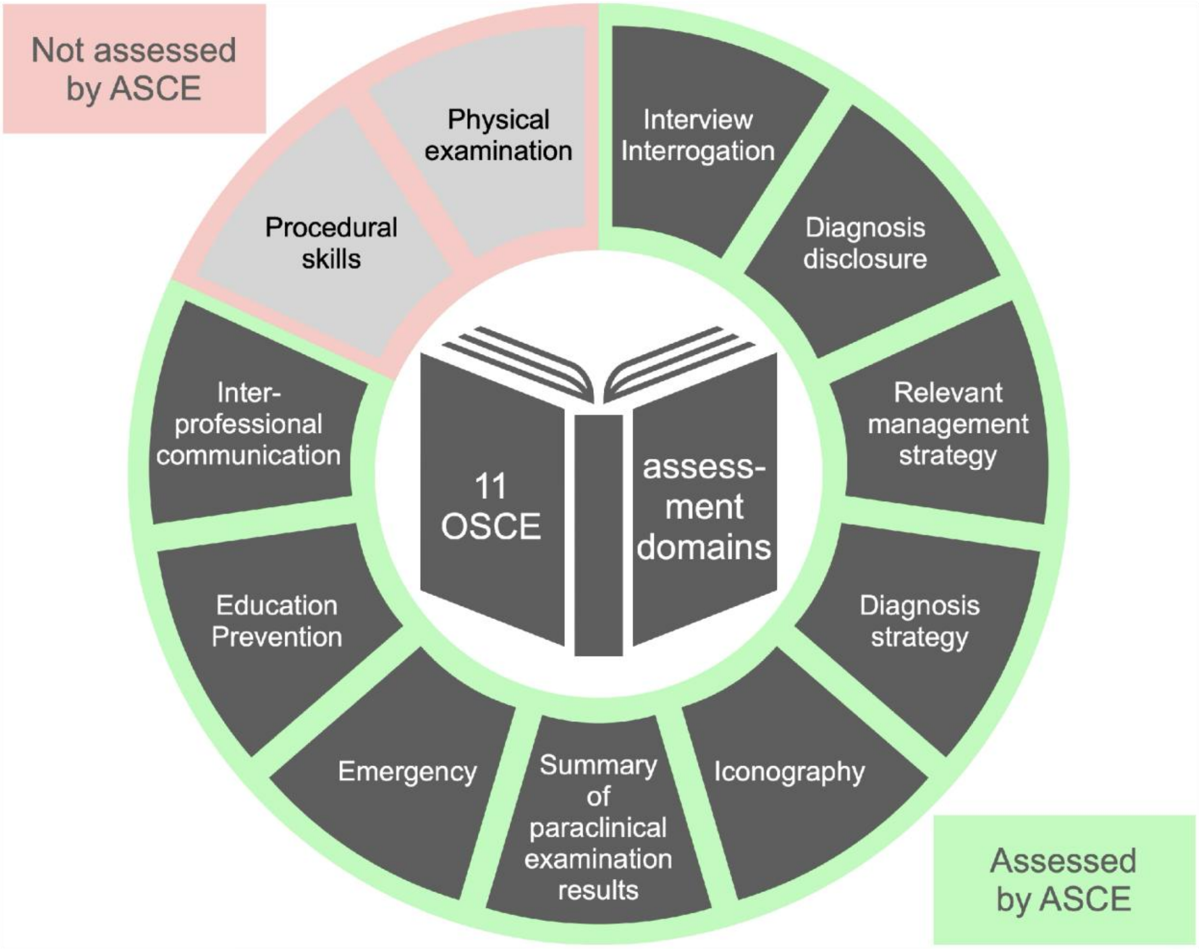

**Figure S2. Selection Bias Analysis.**

Box plots of national OSCE scores among three groups: students who completed all ASCE stations (n = 588), students who connected but did not complete (n = 56), and enrolled students who did not start the ASCE (n = 293). OSCE scores did not differ across groups (Welch analysis of variance, P = 0.66); pairwise comparisons using the Games-Howell post-hoc test likewise showed no significant differences (all P ≥ 0.64). Students who were eligible but never enrolled had no national score and could not be compared. ASCE denotes AI-standardized clinical examination, and OSCE objective structured clinical examination.

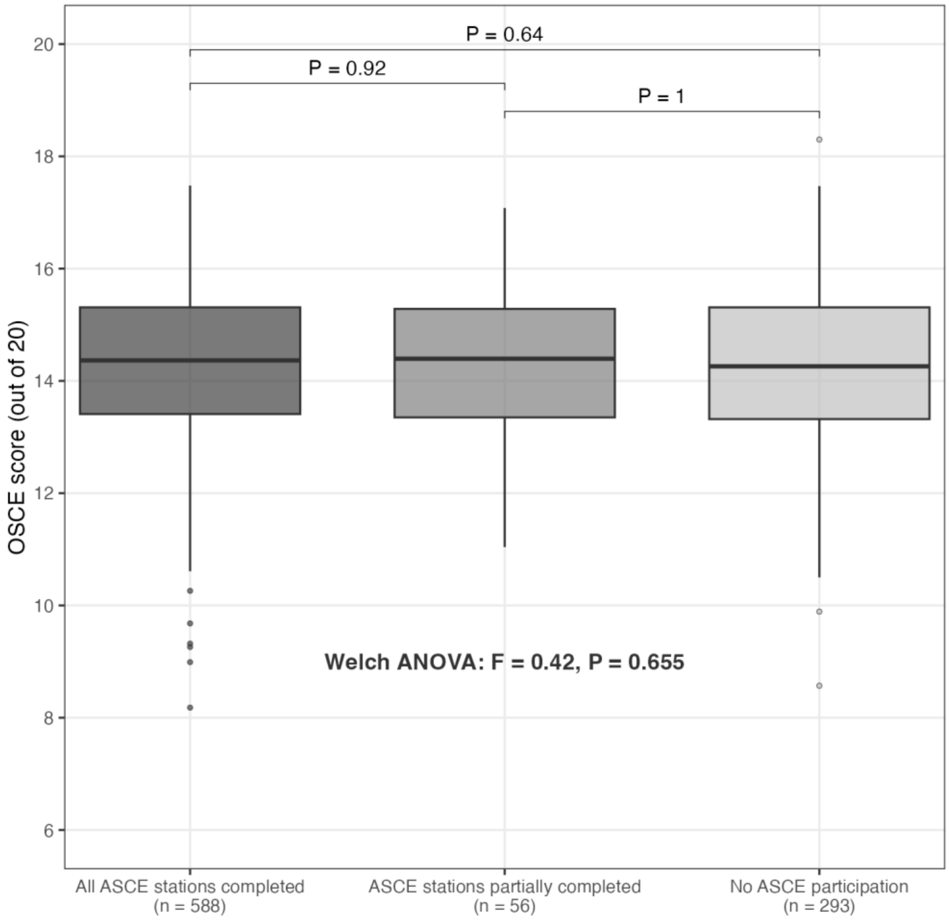

**Figure S3. Distribution of ASCE and OSCE Scores.**

Overlaid histograms of ASCE scores among the 589 students who completed all 10 stations (mean 10.87 ± 1.47) and national OSCE scores among the 588 of them with a national score (mean 14.26 ± 1.45), both on a 20-point scale, with the two axes aligned on their medians. Both distributions are unimodal with a mild left tail; ASCE scores are centered 3.4 points lower than national OSCE scores. ASCE denotes AI-standardized clinical examination, and OSCE objective structured clinical examination.

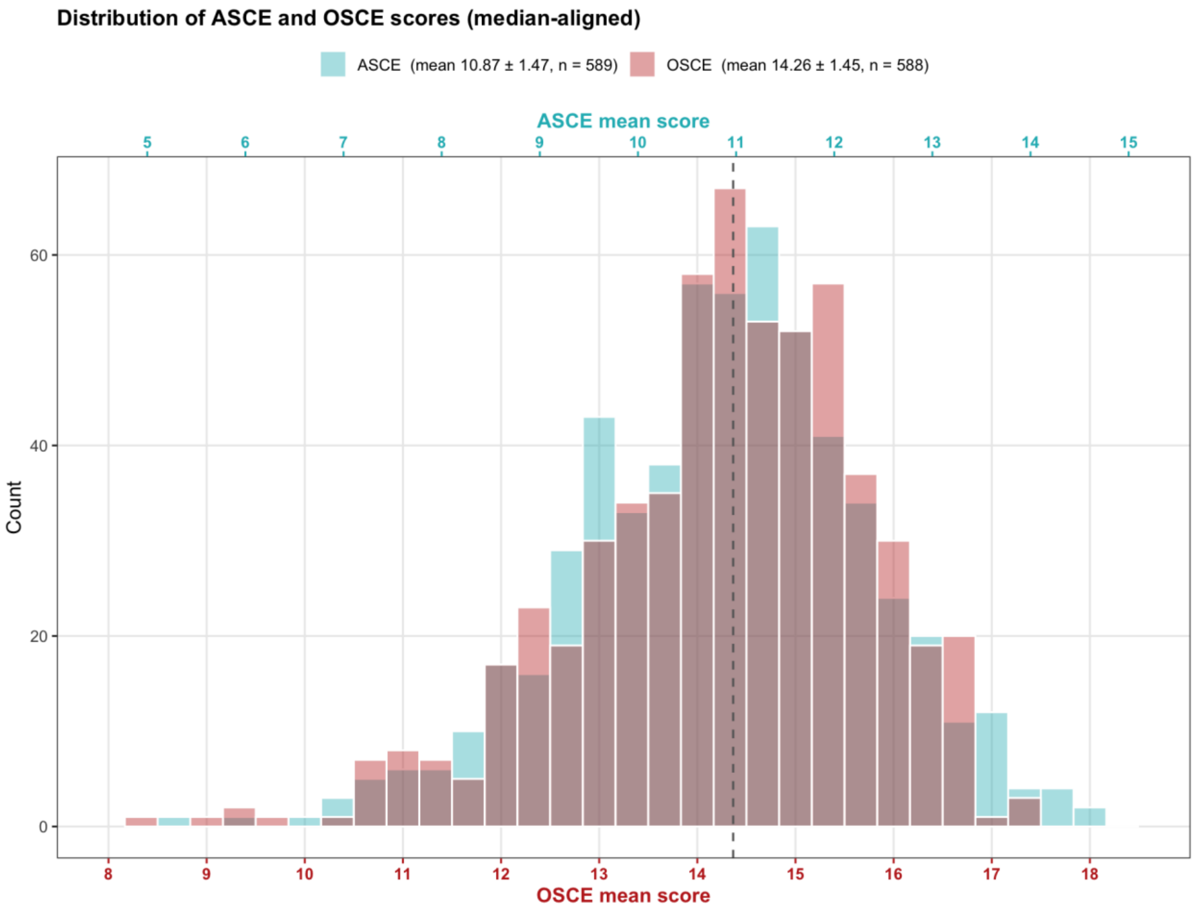

**Figure S4. Clinical Skills Scoring by Evaluator.**

Violin plots showing the distribution of scoring errors for clinical skills items for each expert evaluator (H1 through H8) and the GPT-4.1 model. The error for each evaluator is calculated as the difference between the individual score and the consensus score derived from the remaining expert evaluators using a leave-one-out methodology; GPT-4.1 was compared with the median of all experts who scored that transcript. The analysis comprises evaluations across 20 distinct ASCE transcripts. The width of each violin corresponds to the frequency of data points at that error magnitude. Evaluators are ordered by increasing median error. ASCE denotes AI-standardized clinical examination.

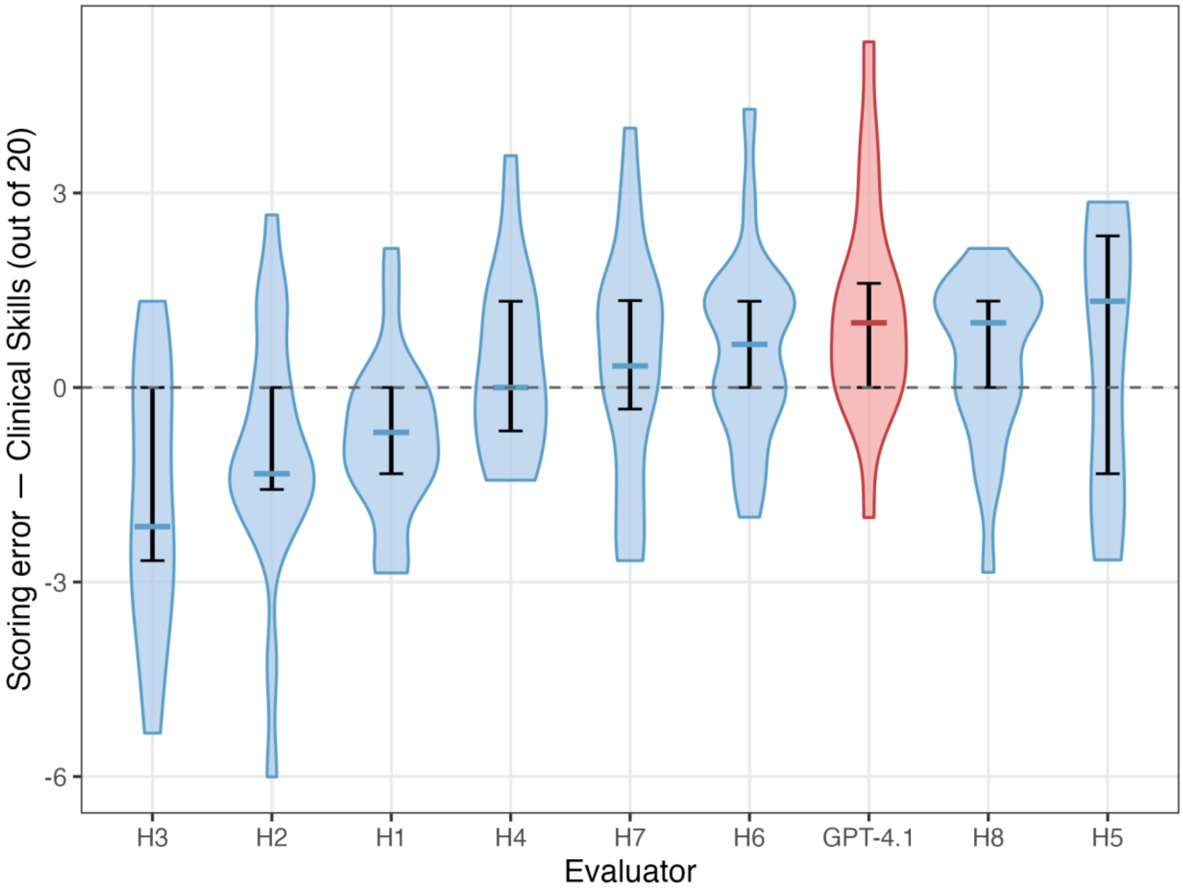

**Figure S5. Communication and Attitude Scoring by Evaluator.**

Violin plots showing the distribution of scoring errors for communication and attitude items for each expert evaluator (H1 through H8) and the GPT-4.1 model. The error for each evaluator is calculated as the difference between the individual score and the consensus score derived from the remaining expert evaluators using a leave-one-out methodology; GPT-4.1 was compared with the median of all experts who scored that transcript. The analysis comprises evaluations across 20 distinct ASCE transcripts. The width of each violin corresponds to the frequency of data points at that error magnitude. Evaluators are ordered by increasing median error. ASCE denotes AI-standardized clinical examination.

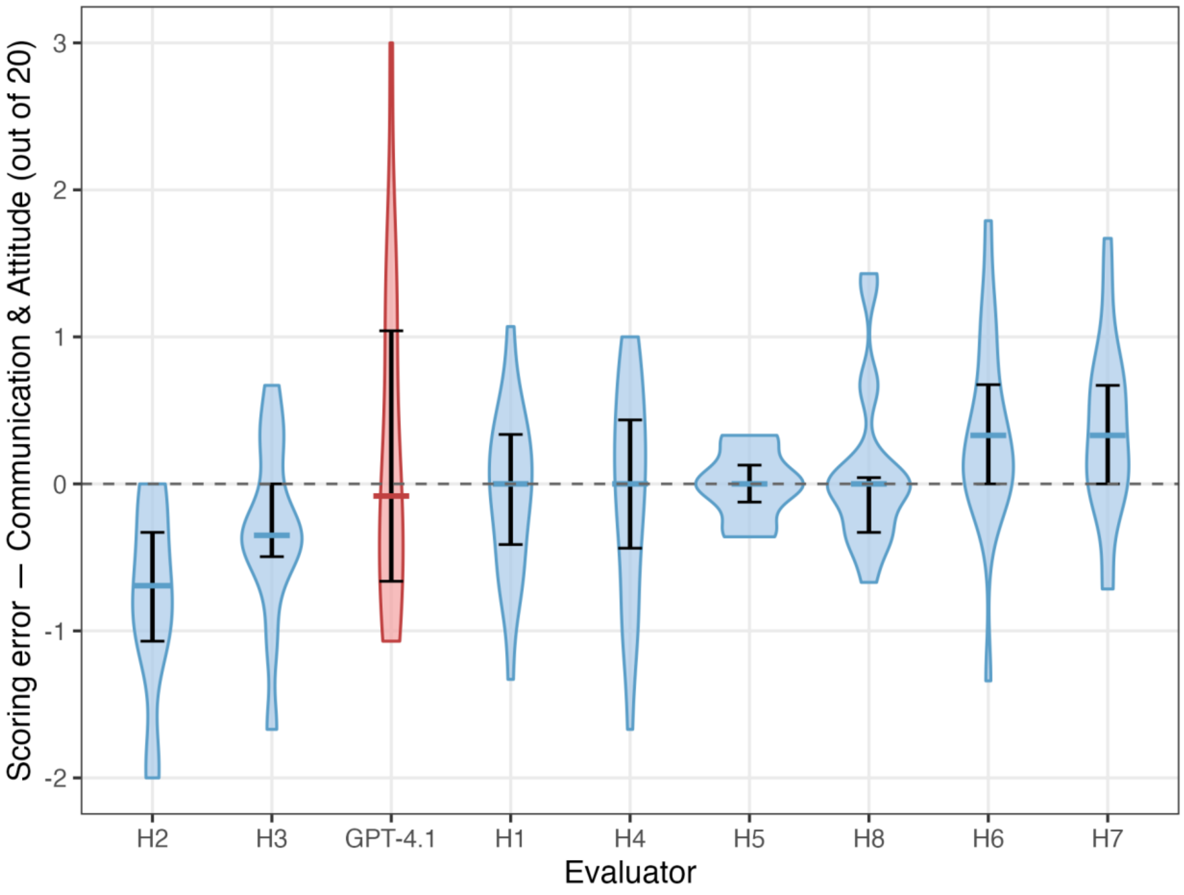

**Figure S6. Predicted Reliability of ASCE.**

The curve illustrates the projected increase in internal consistency (Cronbach's alpha) as a function of the number of ASCE stations. The projection is based on the Spearman-Brown prediction formula applied to the observed data (Cronbach α = 0.67 for 10 stations). Horizontal dashed lines indicate reliability values of 0.7, 0.8, and 0.9. Approximately 12 stations would yield α ≈ 0.70, 20 stations α ≈ 0.80, 30 stations α ≈ 0.86, and 45 stations α ≈ 0.90. ASCE denotes AI-standardized clinical examination.

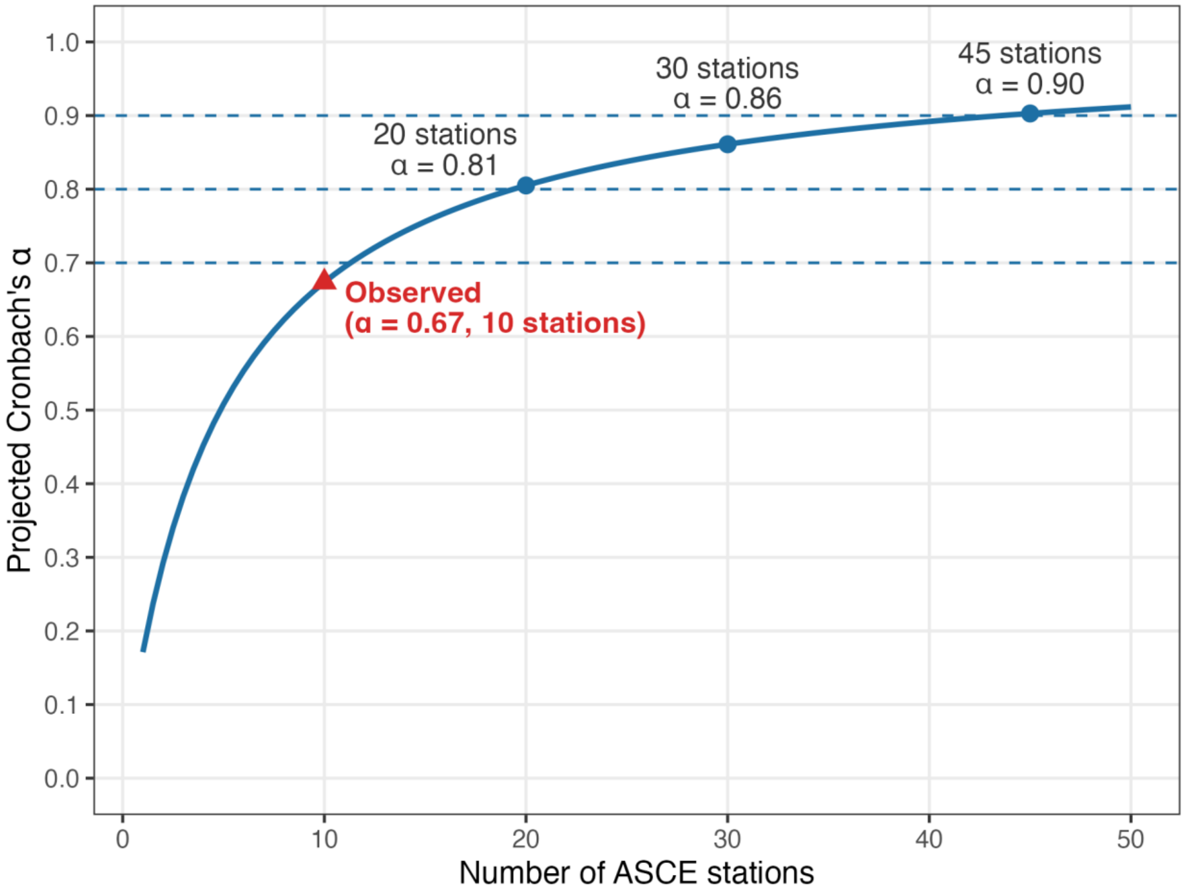
